# Healthcare Workers’ Mental Health and Wellbeing During the COVID-19 Pandemic in the UK: Contrasting Guidelines with Experiences in Practice

**DOI:** 10.1101/2020.07.21.20156711

**Authors:** Norha Vera San Juan, David Aceituno, Nehla Djellouli, Kirsi Sumray, Nina Regenold, Aron Syversen, Sophie Mulcahy Symmons, Anna Dowrick, Lucy Mitchinson, Georgina Singleton, Cecilia Vindrola-Padros

## Abstract

**Background:** Substantial evidence has highlighted the importance of considering healthcare workers’ (HCW) mental health during the COVID-19 pandemic, and several organisations have issued guidelines with recommendations. However, the definition of wellbeing and the evidence-base behind such guidelines remains unclear.

**Objectives:** Assessing the applicability of wellbeing guidelines in practice; identifying unaddressed HCWs’ needs; and providing recommendations for supporting frontline staff during the current and future pandemics.

**Methods and Design:** This paper discusses the findings of a qualitative study based on interviews with frontline healthcare staff in the UK and examines them in relation to a rapid review of wellbeing guidelines developed in response to the COVID-19 pandemic.

**Results:** 14 guidelines were included in the rapid review and 33 interviews with HCWs were conducted in the qualitative study. As a whole, the guidelines placed greater emphasis on wellbeing at an individual level, while HCWs placed greater emphasis on structural conditions at work, such as understaffing and the invaluable support of the community. This in turn had implications for the focus of wellbeing intervention strategies; staff reported an increased availability of formal mental health support, however, understaffing or clashing schedules prevented them from participating in these activities.

**Conclusion:** HCWs expressed wellbeing needs which align with social-ecological conceptualisations of wellbeing related to quality of life. This approach to wellbeing has been highlighted in literature about HCWs support in previous health emergencies, yet it has not been monitored during this pandemic. Wellbeing guidelines should explore staff’s needs and contextual characteristics affecting the implementation of recommendations.

## 1 Introduction

On 11 March 2020, the World Health Organization (WHO) declared COVID-19 a global pandemic [1]. The rapid spread of this virus, with limited effective treatments currently available, has led to a global crisis with overwhelmed health systems that have had to adapt rapidly to respond to the emergency. This has particularly been the case for the UK, the worst affected country in Europe, where at the time of writing at least 286,194 people have been infected and 40,542 have died [2].

Healthcare systems around the world have been under high demand, with frontline staff exposed to unprecedented strain. Healthcare workers (HCWs) have been facing work overload, fear of infection, frustration, discrimination, isolation and lack of contact with their families [3]. Substantial evidence from similar extreme situations, including previous epidemics, has highlighted the importance of considering frontline workers’ mental health and wellbeing [4]. HCWs work long hours under pressure, often without proper resources and facing inherent dangers and lack of clarity regarding the limits of their duty of care [5]. Several organisations have issued guidelines with recommendations on how to protect healthcare workers’ wellbeing [6]. However, the definition of wellbeing and the evidence-base behind such guidelines remains unclear. There is a tension in the wellbeing literature between individual/clinical and ecological approaches, which in turn have implications for the focus of interventions [7]. Critics of the clinical model of wellbeing have highlighted limitations regarding the lack of sensitivity across contexts and not including outcomes that are meaningful to end-users [8]. Consequently, proposed guidelines based on this model may fall short when addressing HCWs’ wellbeing needs in practice and might not be aware of potential barriers in the implementation of recommendations.

This paper discusses the findings of a qualitative study based on interviews with frontline healthcare staff in the UK and examines them in relation to a rapid review of wellbeing guidelines developed in response to the COVID-19 pandemic. The aims of the study are to assess the applicability of guidelines in practice; identify unaddressed HCW needs; and provide recommendations for supporting frontline staff during the current and future pandemics.

## 2 Methods

### 2.1 Rapid synthesis of wellbeing guidelines

#### Search strategy

We conducted a review of the literature following the Preferred Reporting Items for Systematic Reviews and Meta-Analyses (PRISMA) guidelines [9]. In view of the quickly evolving situation we adopted a rapid review methodology, following approaches developed by the WHO and the Cochrane Collaboration for rapid evidence synthesis [10,11]. A protocol was developed before searching and registered in PROSPERO (CRD42020183393).

We searched for articles and guidelines providing recommendations to promote wellbeing, improve mental health or prevent mental health problems in healthcare staff during the COVID-19 pandemic. The search strategy included terms such as “wellbeing”, “mental health”, “coping”, “healthcare workers” and “covid”. Searches were conducted in PubMed, EMBASE and PsycInfo, and grey literature searches through OpenGrey and Tripdatabase. The search was conducted on the 23^rd^ of April. Additionally, we hand-searched key websites and online databases of government institutions and professional societies in the UK to identify guidelines that may not have been published elsewhere.

Finally, we supplemented our search by cross-referencing the included studies. We included articles written in English, with a focus on the UK setting, although international recommendations were included due to their potential influence on local guidelines and practice. A full description of the search strategy can be found in the Supplementary material 1.

#### Study selection, data extraction and risk of bias assessment

To maintain consistency throughout the study selection, three team members (DA, DJ, SMS) independently screened titles and abstracts and then assessed whole texts of eligible articles against the full review inclusion and exclusion criteria. We extracted relevant information from the included articles alongside wellbeing recommendations, such as date, specific healthcare staff groups, conceptual framework and evidence supporting recommendations. We synthesised the extracted data based on recommendations to maintain wellbeing and prevent mental illness in an aggregative/descriptive manner, summarising information into categories that were common across the included guidelines. Finally, the Reporting Items for Practice Guidelines in Healthcare (RIGHT) tool [12] was used to appraise the methodological quality of the included guidelines.

### 2.2 Qualitative study: frontline staff perceptions and experiences of wellbeing

#### Data collection

This qualitative study is part of a larger ongoing project conducted by the Rapid Research, Evaluation and Appraisal Lab (RREAL) which was designed as a qualitative rapid appraisal combining three streams of work: policy review, media and social media analysis, and telephone interviews [13]. Rapid appraisals are developed to collect and analyse data in a targeted and iterative way within limited timeframes, and ‘diagnose’ a situation [14,15].

A purposive sample of HCWs was selected for interview based on their role in 15 health facilities in the UK. In this paper, we report on the findings from the first 33 interviews carried out with frontline staff between March 19^th^ and April 24^th^, 2020.

Semi-structured interviews were carried out by KS, AS, LM, GS and CV. Interviewers received specific training on data security awareness from NHS Digital and Health Education England. There was no relationship between interviewers and interviewees prior to the interviews and correspondence was limited to arranging a time for the interview to take place. Interviews were conducted over the phone, audio-recorded and additional notes on the main topics discussed were documented (see the interview topic guide in Supplementary material 2).

All personal identifiers from the interview transcripts were removed. Data were kept on a secure server and interviewees were grouped in generic role categories, rather than job title, to avoid individuals being identifiable when quoting from interviews.

The authors assert that all procedures contributing to this work comply with the ethical standards of the relevant national and institutional committees on human experimentation and with the Helsinki Declaration of 1975, as revised in 2008. This study was approved by the Health Research Authority (HRA) in the UK (IRAS: 282069) and the local Research and Development Offices (R&D) where the study took place. All participants provided consent before taking part.

A table summarising the three workstreams and sample of the project as a whole can be found in the Supplementary material 3.

#### Data extraction and analysis

A group of the authors (NV, KS, ND, NR, AS) performed selective transcription of extracts from the interviews and interview notes that were related to mental health and wellbeing. We also included additional information on demographic characteristics of the participants to contextualise the information discussed in the transcripts.

We analysed the data using framework analysis [16]. We developed an analytical coding framework based on a preliminary scan of the data and inputted it in a Microsoft Excel matrix, with the emerging codes in the columns and cases in rows. The framework was refined during team discussions and we used the final set of agreed codes to index all the interview data. After indexing was completed, we synthesised the key topics emerging within each code and, based on this, developed the final set of themes encompassing the main issues raised by frontline staff. The team also selected quotes from the interview transcripts that could exemplify these themes.

## 3 Results

### 3.1 Synthesis of wellbeing guidelines

We identified 255 unduplicated papers/reports from databases and additional sources. After screening titles and abstracts, 21 articles were reviewed in full text. A final 14 articles were included in this review according to the pre-specified inclusion criteria. The PRISMA flowchart showing this process is depicted in Figure 1.

**Figure 1.**
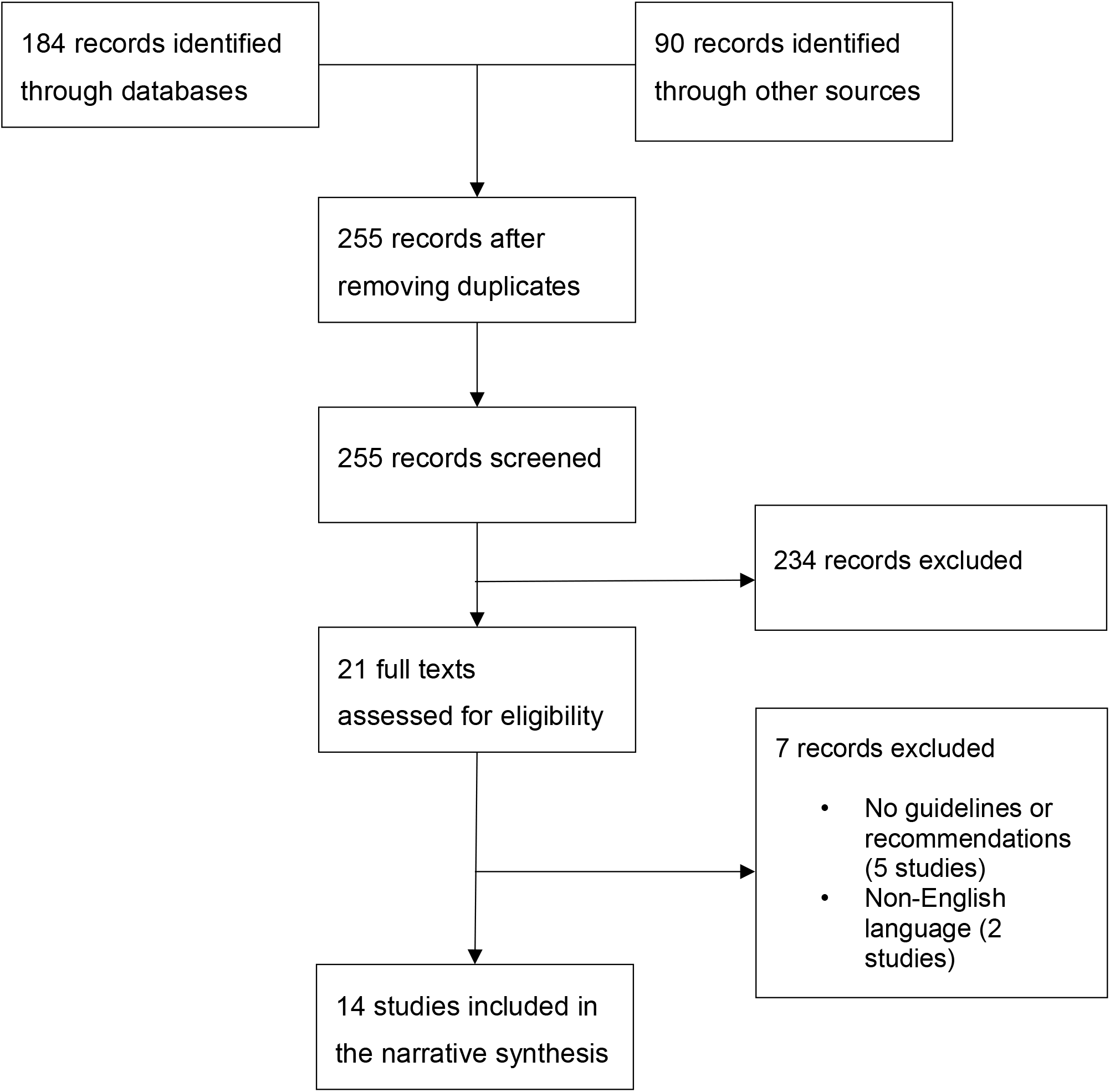
PRISMA flowchart.

The main characteristics of the included studies can be found in Supplementary material 4. The studies were published at the peak of the UK pandemic, between March and April 2020. Nine articles consisted of practical recommendations published on institutional webpages and local repositories, and five were journal articles. Guidelines proposed by institutions were developed mostly for the attention of healthcare managers and leaders, while the five journal articles were aimed at giving wellbeing and mental health recommendations directly to HCWs delivering care to COVID-19 patients.

Three reports [17–19] highlighted that work under pressure and risk of burnout were pre-existing conditions among HCWs. As a result, the guidelines highlighted self-monitoring and help-seeking to prevent more severe mental health problems and recommended managers to be proactive in detecting mental health concerns among staff and providing psychological support. We did not find any recommendation to use specific interventions or screening tools to detect mental illness.

The guidelines’ theoretical assumptions around the concept of wellbeing and how this is affected by the pandemic were stated in 10 articles. One assumption regarding wellbeing was based on a psychosocial resilience approach [17], referencing a framework which focused on individuals maintaining social ties through online platforms and managers reinforcing team working [20]. Nine guidelines based their recommendations on the individual’s potential to develop mental illness, post-traumatic stress disorder (PTSD) in particular, and individual needs emerging from exposure to the virus or difficult life and death decisions. Four guidelines did not specify any conceptual framework or definition of wellbeing to inform their recommendations.

Explicit and non-explicit theoretical assumptions influenced the types of recommendations made to support HCWs. We found that recommendations regarding wellbeing corresponded to three overarching categories: institutional/organisational level recommendations; individual level recommendations; and recommended resources for further help. Organisational-level recommendations focused on the importance of leadership, team communication, peer support, flexibility in working shift patterns and providing psychological support to HCWs. At an individual level, the guidelines highlighted the role of trauma and response to mental health crisis; the importance of allocating time for healthy eating and sleeping habits; attending relevant clinical training to improve self-efficacy and an individual’s sense of control; keeping in touch with support networks; and access to psychological therapy and adequate personal protective equipment (PPE). Three guidelines suggested psychological and peer debriefing; however, this was discouraged by another guideline [21]. Lastly, practical advice included limiting the consumption of news and social media to avoid feeling overwhelmed by information about the pandemic, the practice of relaxation and mindfulness techniques, and setting up rapid ethics boards to support difficult end-of-life decisions.

A detailed list of the recommendations from the guidelines can be found in the Supplementary material 5.

The quality appraisal based on the RIGHT tool yielded heterogeneous results. Most guidelines were clear about the aims, the target population and the context where the recommendations ought to be applied. Wellbeing recommendations were generally precise and formulated in a manner that allowed the development of actionable measures. The guidelines provided little detail of the evidence behind their suggestions and none of them specified the certainty or strength of their recommendations. A detailed analysis with the RIGHT tool can be found in Supplementary material 6.

### 3.2 Qualitative study: frontline staff perceptions and experiences

#### Sample

We conducted interviews with 20 female HCWs and 13 male HCWs with various roles (nurses, doctors with different specialties including anaesthetists, surgeons and GPs, healthcare assistants and pharmacists). Additionally, our interview sample included a wide range of work experience, from recently qualified HCWs to HCWs with almost 40 years of experience in the NHS.

#### Themes

Five themes emerged from the qualitative interviews: *Wellbeing and “pulling together”*; *Concerns, unsettling experiences and difficult moments*; *Experiences around PPE*; *Morale and barriers to performing confidently*; *and Life outside the clinical role*. The text below defines each theme and key quotes are presented in Table 1. The Supplementary material 7 provides an overview of key characteristics and additional illustrative quotes for each theme.

**Table 1.**
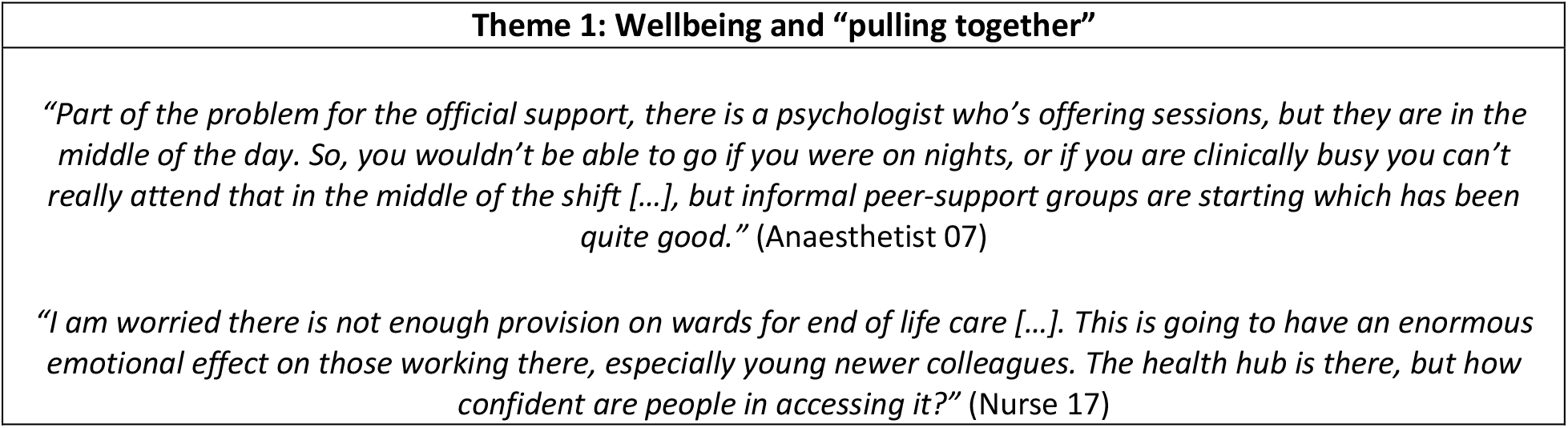
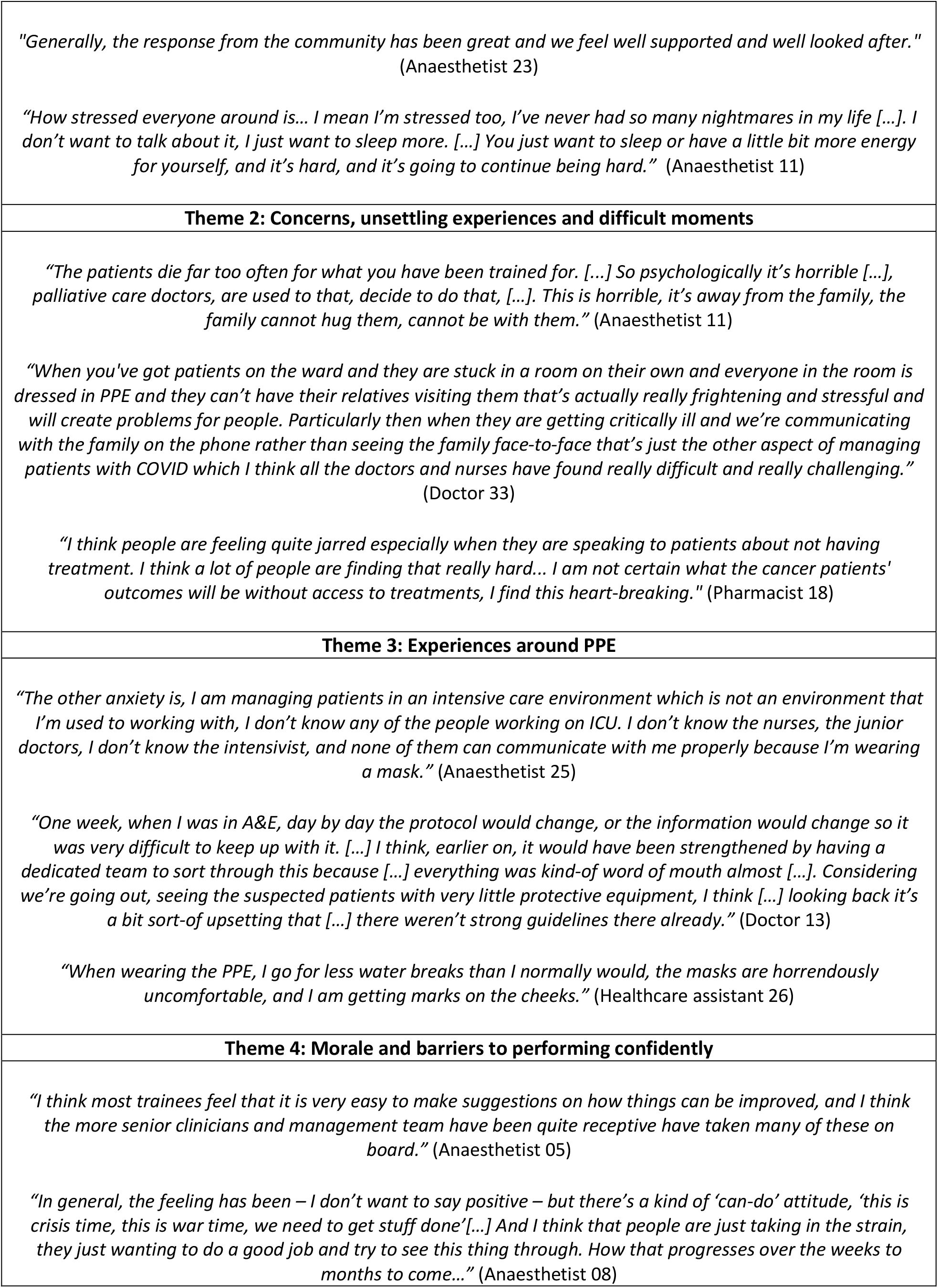
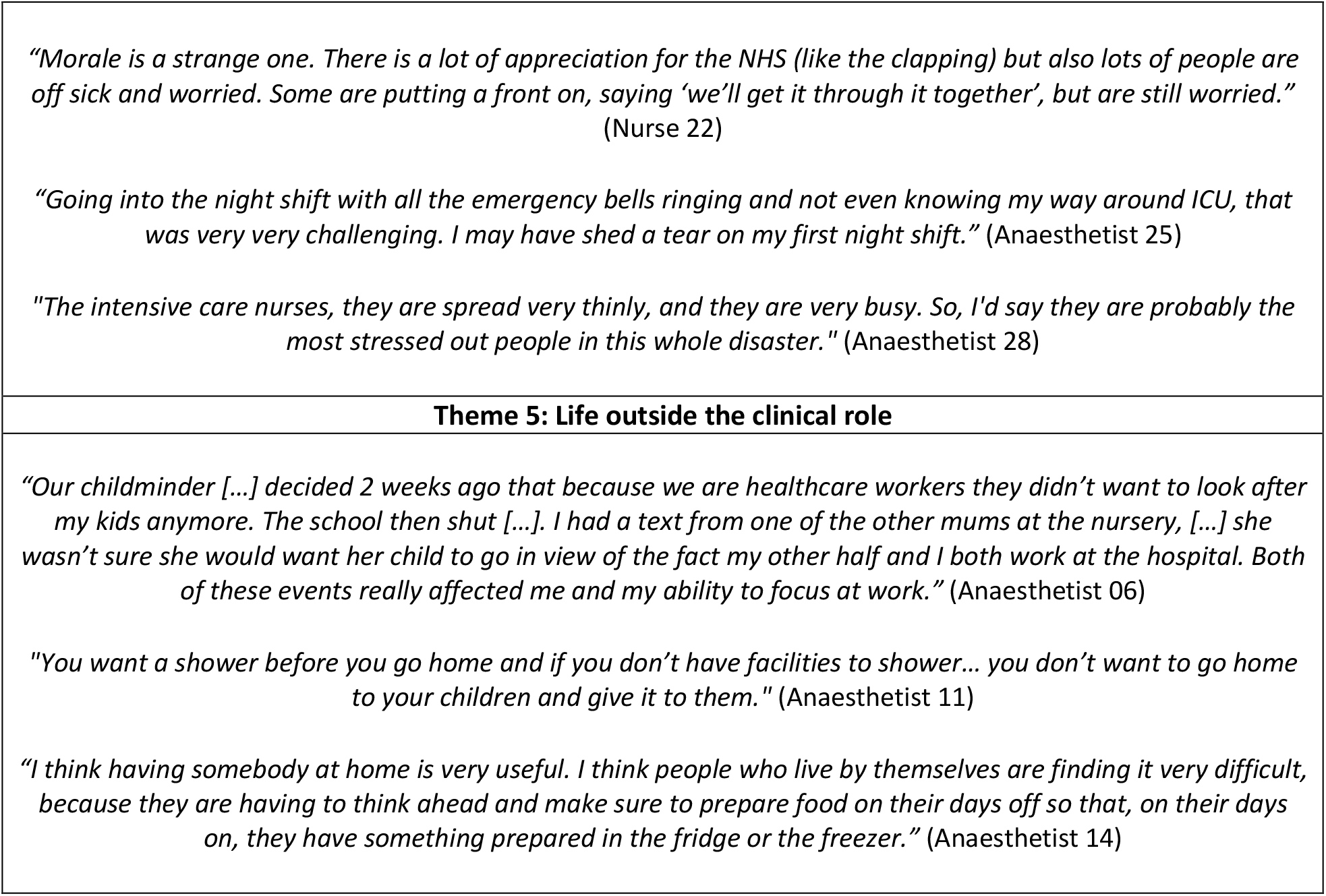
Key participant quotes illustrating the qualitative findings.

#### Wellbeing support and “pulling together”

There was a significant focus on HCWs’ mental health and psychological support as part of service adaptations during the pandemic. Examples of this were increasing the availability of clinical psychologists to provide therapy, online counselling or group support sessions. However, staff reported that additional workload or clashing schedules prevented them from participating in these formal wellbeing activities. Some staff mentioned there was “*no room”* for wellbeing sessions during the peak of the pandemic and they prioritised resting during breaks.

The most salient form of wellbeing support mentioned by staff was mutual moral and clinical support between HCWs. This support happened through buddy systems and spontaneously as a general feeling of motivation, comradery, and empowerment among teams. Factors that were said to contribute to positive team dynamics were a less hierarchical structure and maintaining consistency in the composition of working teams. Positive group dynamics between colleagues could be negatively affected by the lack of clarity around duty of care between different medical specialties and the lack of testing for HCWs. This last point was considered woefully inadequate and distressing for staff and their families, as well as exacerbating guilt from being off work without a “justified” cause.

HCWs felt appreciated and valued owing to community support, citing examples such as rainbow pictures and clapping. Food donated by local restaurants and neighbours was reported to have a significant impact on keeping morale high and getting staff through long shifts.

#### Concerns, unsettling experiences and key difficult moments

Participants described a range of struggles at different stages of the pandemic. In the pre-peak and preparation stages, HCWs experienced anxiety and anticipation mainly in relation to: news reports of international experiences in Intensive Care Units (ICU); the possibility of bringing the virus home to families; and moving into a “greatest good for the greatest number” model of care (where decisions needed to be made about the rationing of scarce services between patients) that could have moral and legal implications for clinicians. Worries were most alleviated through training, gaining experience in new roles, and transparent and consistent communications from managers. In general, staff reported that anxiety diminished as they became more immersed in their work and, in some cases, once they had contracted and recovered from the virus. Those who worked in particularly busy departments expressed anxiety about understaffing due to sickness when more COVID-19 positive patients started presenting.

During the height of the pandemic, staff mentioned experiencing additional cognitive burdens to their usual work, such as uncertainties around diagnosis without the usual diagnostic techniques available and feeling overwhelmed by frequent changes in PPE and clinical guidelines. At the same time, staff spoke about additional moral labour to their usual ICU work because patients’ families were not allowed to visit. Two moments identified as particularly difficult in this respect: when patients were initially taken into the ICU and in cases where patients were dying. HCWs spoke of difficulties interacting with the families of dying patients remotely.

As the peak passed, concerns shifted towards the backlog of non-COVID-19 patients who had not received care during this period, and fears regarding how staff and patients would cope with a second peak.

#### Experiences around PPE

PPE was a consistent challenge for all HCWs. During the early stages of the pandemic, HCWs experienced anxiety in relation to the correct method of donning and doffing of PPE. Later in the pandemic, working in full PPE and not having proper breaks was mentioned as a factor generating great distress. Staff reported overheating, dehydration, exhaustion and expressed discomfort due to PPE being “*one size fits all*” and thus causing pain or feeling claustrophobic. Rapidly changing and conflicting guidelines on PPE use caused anxiety. Many HCWs worried guidelines were changing because of lack of stock rather than due to safety standards.

Staff emphasised working in full PPE as a factor that hindered communication with colleagues and the generation of a close relationship with patients and their families. In particular, staff mentioned “*everything came out as a shout*” and it was not possible to identify colleagues behind masks.

Staff expressed ambivalence about how families should have been involved in patient care, questioning whether it was appropriate for them to visit patients and wear PPE. The possibility of patients dying alone had generated great distress for HCWs, patients and families, so some HCWs felt family accompanying dying patients should be a priority. Others commented feeling nervous due to large families making inappropriate use of PPE before regulations began.

#### Morale and barriers to performing confidently

Staff reported morale was generally high; they felt positive changes to services had happened efficiently, senior management were responsive, and junior doctors and nurses felt empowered as their suggestions were being heard. However, participants expressed concerns that morale may deteriorate as weeks went by working under strenuous conditions. An important barrier to performing confidently was lack of sleep due to increased workload to cover staff sickness and an increment in the number of night shifts that staff were required to do.

Challenges varied between professionals; junior doctors faced uncertainty regarding exams and courses that had been paused, and consultants were required to manage much larger teams and keep up to date with frequently changing protocols. Regarding the latter, doctors working with COVID-19 patients found their self-efficacy was weakened by lack of clarity regarding clinical protocols.

In particular, ICU nurses were thought to have had the largest work increase and be emotionally affected by not being able to provide the quality of work they were used to. Another significantly affected group were anaesthetists, who had to rapidly take on new roles as intensivists, working alongside new colleagues and under the constraints of PPE.

#### Life outside the clinical role

The professional experiences of HCWs during the pandemic were greatly influenced by their lives outside of work. Some staff reported initially experiencing negative interactions with neighbours or childminders who perceived them to be at greater risk of infection. However, for the most part, participants expressed feeling incredibly fortunate and grateful for the community support.

The main concerns outside the hospital were caring for children and elderly relatives, completing shopping and housework and arranging their travel to and from hospital. Navigating caring duties proved complicated; supportive families and sustained schooling provided key relief. Staff appreciated the senior management team planning to cover needs beyond HCW clinical work such as providing free parking to facilitate travel to and from work, optional accommodation, or hospital childcare remaining available. Also helpful were bicycle and transport providers offering free rides for NHS workers.

Physical spaces such as ‘wobble’ rooms and health hubs were reported to provide much-needed areas to rest and to re-charge. However, working facilities varied across sites. Some staff described a lack of cleaning facilities and showers to prevent contamination and few spaces for resting.

HCWs mentioned the increased difficulty of maintaining a life-work balance due to the combination of increased workload and the limited number of leisure options available. People who lived alone found isolation particularly difficult and some reported being expected to do significantly more clinical work in order to cover for colleagues who had family responsibilities. Isolation was also particularly difficult for people with pre-existing mental health conditions or difficult situations at home.

## 4 Discussion: Mapping guideline themes and staff experiences

Multiple guidelines were available to safeguard and support HCWs as the pandemic reached its peak. In this sense, we can conclude that there was a high awareness of mental health and wellbeing needs. Guidelines as a whole tended to concur with staff’s concerns. However, there was discordance in terms of where emphasis was placed, and important gaps in guidelines in relation to the role of the community support in wellbeing and contextual barriers to materialise recommendations in practice.

As a whole, the guidelines placed greater emphasis on wellbeing at an individual level. This was under the premise that staff were likely to develop mental disorders due to having to make difficult life or death decisions or being exposed to the virus without appropriate PPE. This clinical approach to wellbeing speaks of mental distress in terms of diagnosis and frames wellbeing in terms of outcome measures of symptoms and psychosocial functioning designed by mental health professionals [22,23]. Cross-sectional studies about staff mental health conducted in China during the pandemic, for example, reported percentages of staff experiencing mild to severe anxiety ranging from 23-45% [24,25]. Staff, however, placed greater emphasis on structural conditions at work, such as understaffing and the invaluable support of the community. This aligns with social-ecological conceptualisations of wellbeing related to quality of life, which have been highlighted in literature about HCWs support and wellbeing in health emergencies [26,27], yet they have not been monitored during this pandemic.

The misalignment between what staff perceived as important and what was prioritised in wellbeing guidelines highlights that it is imperative to involve staff at all stages of developing wellbeing support strategies in order to incorporate their input about how to respond to local needs and contexts. This was one of the five key recommendations of the Boorman review, commissioned by the UK Department of Health specifically to address healthcare staff wellbeing. This review, and subsequent research derived from it, advocates for a “whole-system” and participatory approach to create wellbeing programmes [7]. At the same time, the need to take a holistic approach to planning wellbeing aid in emergencies has been highlighted in emergency responses to other epidemics, such as modern approaches to HIV prevention which are directed towards addressing structures that constrain or enable people’s choices [26]. A focus on mental illness, PTSD in particular, has been criticised in past mental health emergency interventions, citing its lack of attention to practical wellbeing needs and potential misconception of healthy individual responses [28–30]. An example of this in our study was a guideline suggesting that staff who repeatedly expressed not being available to attend peer support groups should be interpreted as using “avoidance” and it being a potential symptom of trauma; staff, on the other hand, reported having very few breaks due to understaffing and wishing to dedicate these breaks to resting. Additional ambiguity in interpretations can be expected to arise due to lack of recommendations in guidelines around specific interventions or screening tools to detect mental illness.

Situating practice in its context and combining organisational, community and personal strategies for wellbeing has shown to increase the likelihood of success in wellbeing promotion interventions [31,32]. Findings from this study pointed to the role of community support as being a key contextual facilitator of wellbeing. In this respect, an important shift in community attitudes towards HCWs was identified; staff and their families were initially stigmatised and later admired and rewarded by the community. These findings suggest that implementing “appreciation” and anti-stigma campaigns to promote support to HCWs should be included as a key preparation to support staff’s wellbeing [33].

An organisational factor which stood out in guidelines and the qualitative findings was HCWs’ feelings around being stronger working as a collective and “pulling together” in less hierarchical working conditions, while at the same time maintaining strong leadership and guidance from managers. This is recognised in literature about healthcare staff wellbeing which presents leadership styles focused on transparency, consistency and empowerment of staff lead to higher employee engagement and work satisfaction [32]. However, results from the qualitative study showed senior staff faced great difficulty in keeping up their supporting roles due to having to manage larger teams composed of people they may not have worked with previously. Gaggioli & Riva (2013) propose using apps to monitor individual wellbeing as a strategy that is more flexible and can accommodate staff needs, while at the same time reducing the time required for managers and staff to communicate about this. Further research is needed to ascertain the toll of additional responsibilities on staff performing leadership roles during medical emergencies and potential strategies to address this.

The findings of this study should be considered in light of its strengths and limitations. The rapid review and qualitative study were conducted by a multidisciplinary team, following strong systematic research methods and guidelines, and allowed for a rich discussion contrasting both sources of data to reveal gaps between guidelines and practice. Applicability of the findings outside the context of this study should be evaluated, whilst taking into consideration the rapidly changing circumstances of the current health emergency. New wellbeing guidelines may emerge later in the pandemic or changes affecting HCWs experiences may occur, which we will not be able to capture. Strategies were put in place to maximise rapport during interviews and acknowledge this potential bias at analysis stage. However, sensitive topics, in particular with relation to potential alcohol or drug consumption, were not present in participants’ answers. This should be taken into consideration due to the high relevance of this topic for wellbeing.

To the authors’ knowledge this is the first study contrasting wellbeing guidelines and staff experiences in practice during the COVID-19 emergency. The findings in this study extend the understanding of wellbeing guidelines in the light of HCWs’ experiences in practice. Wellbeing guidelines should take additional steps beyond focusing on top-down clinical understandings of wellbeing, and rather explore staff’s needs and contextual characteristics affecting the implementation of recommendations. Guidelines might need to be developed differently, involving participatory approaches that account for changes in HCWs’ wellbeing over time and allow for the development of responsive and adaptative wellbeing programmes.

## Data Availability

All data relevant to the study are included in the article or uploaded as supplementary information.

## Acknowledgements

We acknowledge the contributions of Dr. Ginger Johnson, who provided general supervision of the larger research project, and Dena Javadi, who provided support with data curation for this study.

## Notes

### Competing Interest Statement

We declare no competing interests.

### Clinical Protocols

https://www.crd.york.ac.uk/prospero/display_record.php?RecordID=183393

### Funding Statement

The study did not receive external funding. Research costs were covered with existing funding provided by the Rapid Research Evaluation and Appraisal Lab (RREAL), UCL. All researchers acted independently from the funding source and can take responsibility for the integrity of the data and the accuracy of the data analysis.

### Author Declarations

This study was approved by the Health Research Authority (HRA) in the UK (IRAS: 282069) and the local Research and Development Offices (R&D) where the study took place. All participants provided consent before taking part.

